# A gut microbiome-derived risk score is associated with future cancer development

**DOI:** 10.64898/2026.09.09.26362619

**Authors:** Emily Oosterhout, Rose S. Leijdesdorff, Oleg Kambur, Bert van der Vegt, Ranko Gacesa, Laura A. Bolte, Marjolein A.Y. Klaassen, Veikko Salomaa, Teemu Niiranen, Rob Knight, Evelien Dekker, Jingyuan Fu, Alexandra Zhernakova, Aki S. Havulinna, Geke A.P. Hospers, Leo Lahti, Rinse K. Weersma, Johannes R. Björk

**Affiliations:** Department of Gastroenterology and Hepatology, University of Groningen and University Medical Center Groningen, Groningen, the Netherlands; Department of Medical Oncology, University of Groningen and University Medical Center Groningen, Groningen, the Netherlands; Department of Gastroenterology and Hepatology, Amsterdam University Medical Center, University of Amsterdam, Amsterdam, the Netherlands; Amsterdam Gastroenterology, Endocrinology and Metabolism, Amsterdam, the Netherlands; Cancer Center Amsterdam, Amsterdam University Medical Center, Amsterdam, the Netherlands; Oncode Institute, Amsterdam, the Netherlands; Institute of Dentistry, School of Medicine, University of Eastern Finland, Kuopio, Finland; Department of Internal Medicine, University of Turku, Turku, Finland; Department of Pharmacology, Faculty of Medicine, University of Helsinki, Helsinki, Finland; Department of Pathology, University Medical Center Groningen, University of Groningen, Groningen, the Netherlands; Department of Pediatrics, University of California San Diego, La Jolla, CA, USA; Center for Microbiome Innovation, University of California San Diego, La Jolla, CA, USA; Department of Computer Science and Engineering, University of California San Diego, La Jolla, CA, USA; Shu Chien-Gene Lay Department of Bioengineering, University of California San Diego, La Jolla, CA, USA; Halıcıoğlu Data Science Institute, University of California San Diego, La Jolla, CA, USA; Hong Kong University of Science and Technology Jockey Club Institute for Advanced Study, Hong Kong University of Science and Technology, Hong Kong SAR, China; Department of Genetics, University of Groningen and University Medical Center Groningen, 9713 GZ Groningen, the Netherlands; Department of Pediatrics, University of Groningen and University Medical Center Groningen, 9713 GZ Groningen, the Netherlands; Institute for Molecular Medicine Finland (FIMM), HiLIFE, University of Helsinki, Helsinki, Finland; Department of Computing, Faculty of Technology, University of Turku, Turku, Finland

## Abstract

Cancer remains a leading cause of morbidity and mortality worldwide, and the incidence of new cancer cases is rising. While established lifestyle factors such as smoking, obesity, and diet contribute to cancer risk, the gut microbiome has emerged as a hallmark of cancer. However, the role of the gut microbiome in cancer has mainly been established based on case-control studies of individuals with cancer at the time of microbiome sampling (prevalent cancer) and through functional studies. Here, we investigated whether the gut microbiome could identify individuals at increased risk of being diagnosed with cancer during follow-up (incident cancer), potentially many years after microbiome sampling. Using shotgun metagenomic sequencing of stool samples from 5,997 participants in the population-based Lifelines cohort and the Dutch Microbiome Project linked to longitudinal cancer registry data, we developed a microbiome-derived risk score based on 28 microbial species. Individuals with higher microbiome risk scores had a higher risk of receiving a cancer diagnosis during up to 10 years of follow-up (adjusted HR per 1-SD increase, 1.62; 95% CI, 1.44-1.82; P=1.54e-15), after adjusting for established cancer risk factors. In independent external datasets comprising 1,879 patients with cancer at the time of microbiome sampling and 5,341 cancer-free controls, the microbiome risk score also distinguished cancer cases from controls. In the independent population-based FINRISK cohort, the score further showed a consistent trend toward reduced cancer-free survival in 328 individuals with incident cancer and 5,636 individuals without cancer during follow-up. Together, these findings demonstrate that the gut microbiome harbors detectable signatures years before cancer diagnosis and highlight its potential as a biomarker for early cancer risk stratification and its potential role in cancer development.

## Main

One in five people are diagnosed with cancer during their lifetime and the annual number of new cases is projected to rise by 77% between 2022 and 2050^1^. Within this global context, Europe accounts for 22.4% of cancer cases and 20.4% cancer-related deaths despite representing less than 10% of the world’s population^1^. Part of this high burden is attributable to a growing and aging global population and increased cancer detection resulting from expanded screening programs^2^. Alongside these trends, the incidence of early-onset cancer, diagnosed before the age of 50, is on the rise in many countries, and has been suggested to reflect multigenerational changes in early-life exposures^3,4^. Lifestyle-related factors, including smoking^5^, alcohol consumption^6^, obesity^7^, and dietary patterns rich in ultra-processed foods^8^, are also increasingly recognized as important modifiable contributors to cancer risk^8,9^. Many of these factors have been associated with alterations of the gut microbiome and disruption of intestinal homeostasis, commonly referred to as gut dysbiosis^10^. The gut microbiome plays a central role in regulating host immunity, metabolism, and maintenance of the intestinal barrier. Consequently, gut dysbiosis has increasingly been implicated in processes linked to tumorigenesis, including chronic inflammation, immune dysregulation, and genotoxic stress, leading to its recognition as a hallmark of cancer^11,12^.

To date, only a small number of microorganisms have been classified as carcinogenic or potentially carcinogenic to humans, most notably *Helicobacter pylori*, which is causally linked to gastric cancer^13^. Most subsequent research has focused on colorectal cancer, for which gut dysbiosis has been consistently observed^14^, and specific bacterial taxa—including *Fusobacterium nucleatum*, colibactin-producing *Escherichia coli*, and enterotoxigenic *Bacteroides fragilis*—have been implicated in pathogenic mechanisms related to colorectal carcinogenesis^15,16^. More recently, this research has expanded beyond gastrointestinal malignancies, with studies suggesting that the gut microbiome may influence the initiation, progression, and treatment outcomes of anatomically distant cancers^17^. Distinct gut microbial signatures have been reported in patients with established lung cancer^18^, breast cancer^19^, prostate cancer^20^, and melanoma^21,22^, while a shared gut microbial signature across multiple cancer types suggests that some microbial alterations may be common to different malignancies^23^. However, because this evidence is derived predominantly from individuals with cancer at the time of microbiome sampling, it remains unclear whether these alterations precede cancer or arise as consequences of the disease or its associated changes. Addressing this question requires large, prospective, population-based cohorts with long-term follow-up to determine whether gut microbial alterations are detectable before a subsequent cancer diagnosis.

We therefore investigated whether the gut microbiome sampled before cancer diagnosis is associated with the risk of a subsequent cancer diagnosis, hereafter referred to as incident cancer, across different cancer types. We analyzed 5,997 adults from the Lifelines biobank with stool shotgun metagenomic profiles generated within the Dutch Microbiome Project (DMP)^10^ and linked these data to histologically confirmed cancer diagnoses from the Dutch NationwidePathology Databank (Palga)^24^. Cancer diagnoses occurring before stool sampling, premalignant or benign lesions, uncertain cancer diagnoses and self-reported previous cancer without a corresponding PALGA record were excluded (**Fig. S1)**. Participants were followed from the day of stool sampling (between January 2013 to September 2016) until cancer diagnosis or the end of available Palga follow-up (16 March 2023), resulting in individual follow-up periods of up to 10 years. During follow-up, 245 participants developed incident cancer and 5,752 remained cancer-free. The most common cancer types were breast (n=58), gastrointestinal (n=46), prostate (n=45), and urinary tract cancers (n=22), together accounting for approximately 70% of all incident cases (**Fig. S2**).

We constructed a microbiome-derived cancer-risk score using an elastic net penalized Cox proportional hazards model applied to the presence-absence profiles of species-level genome bins (SGBs) with Monte Carlo cross-validation comprising 100 repeated random 70/30 train/test splits (**see Methods**). Final feature selection was based on stability across resampling iterations, retaining SGBs with non-zero coefficients and consistent effect directions in at least 90% of resampling iterations. Final risk score coefficients were computed by averaging feature coefficients across all iterations. The final score comprised 28 SGBs whose presence-absence profiles were associated with incident cancer risk (**Table S1**).

In a multivariable Cox regression adjusted for established risk factors such as age, sex, BMI and smoking history, as well as prevalent disease status^10^, Bristol Stool Scale, and the Lifelines diet score, a higher microbiome cancer-risk score was associated with a higher risk of incident cancer diagnosis (HR per 1-SD increase, 1.62; 95% CI, 1.44-1.82; P=1.54e-15, **Fig. 1A**). Consistent with this finding, individuals who were subsequently diagnosed with cancer had higher microbiome cancer-risk scores than those who remained cancer-free during follow-up (P=6.58e-23; **Fig. 1B**). Established risk factors showed associations in the expected direction, including age (HR=1.056, P=1.17e-16) and smoking (HR=1.288, P=0.051). In a sensitivity analysis additionally adjusting for alcohol consumption, 102 participants with missing alcohol data were excluded, leaving 5,895 individuals, including 239 with incident cancer. The association between the microbiome cancer-risk score and incident cancer risk remained similar (HR per 1-SD increase, 1.64; 95% CI 1.45-1.85; P=5.56e-16). High alcohol consumption was also associated with a higher risk of incident cancer diagnosis compared with low alcohol consumption (HR 1.42, 95% CI 1.05-1.93; P=0.0247). The highest microbiome cancer-risk scores were observed among individuals who were subsequently diagnosed with respiratory, skin, male reproductive, and gastrointestinal cancers (**Fig. 1C**).

**Table 1.** Cohort characteristics at stool sample collection.

| **Characteristic** | **Control**<br>N = 5,752<sup>1</sup> | **Case**<br>N = 245<sup>1</sup> | **p-value**<sup>2</sup> | **% Missing** |
| --- | --- | --- | --- | --- |
| **Sex** |  |  | 0.3 | 0.000 |
| Female | 3,407 / 5,752 (59%) | 136 / 245 (56%) |  | 0.000 |
| Male | 2,345 / 5,752 (41%) | 109 / 245 (44%) |  | 0.000 |
| **Age** | 50 (11) | 58 (10) | <0.001 | 0.000 |
| **BMI** | 26.0 (4.2) | 26.5 (3.8) | 0.014 | 0.000 |
| **Prevalent disease** | 4,667 / 5,752 (81%) | 207 / 245 (84%) | 0.2 | 0.000 |
| **Ever smoking** | 2,084 / 5,752 (36%) | 117 / 245 (48%) | <0.001 | 0.000 |
| **Bristol Stool Chart** | 3.79 (0.88) | 3.80 (0.96) | 0.5 | 0.317 |
| **LifeLines diet score** | 25 (6) | 26 (6) | 0.030 | 2.435 |

<sup>1</sup> n / N (%); Mean (SD)<sup>2</sup> Pearson's Chi-squared test; Wilcoxon rank sum test

**Figure 1.**
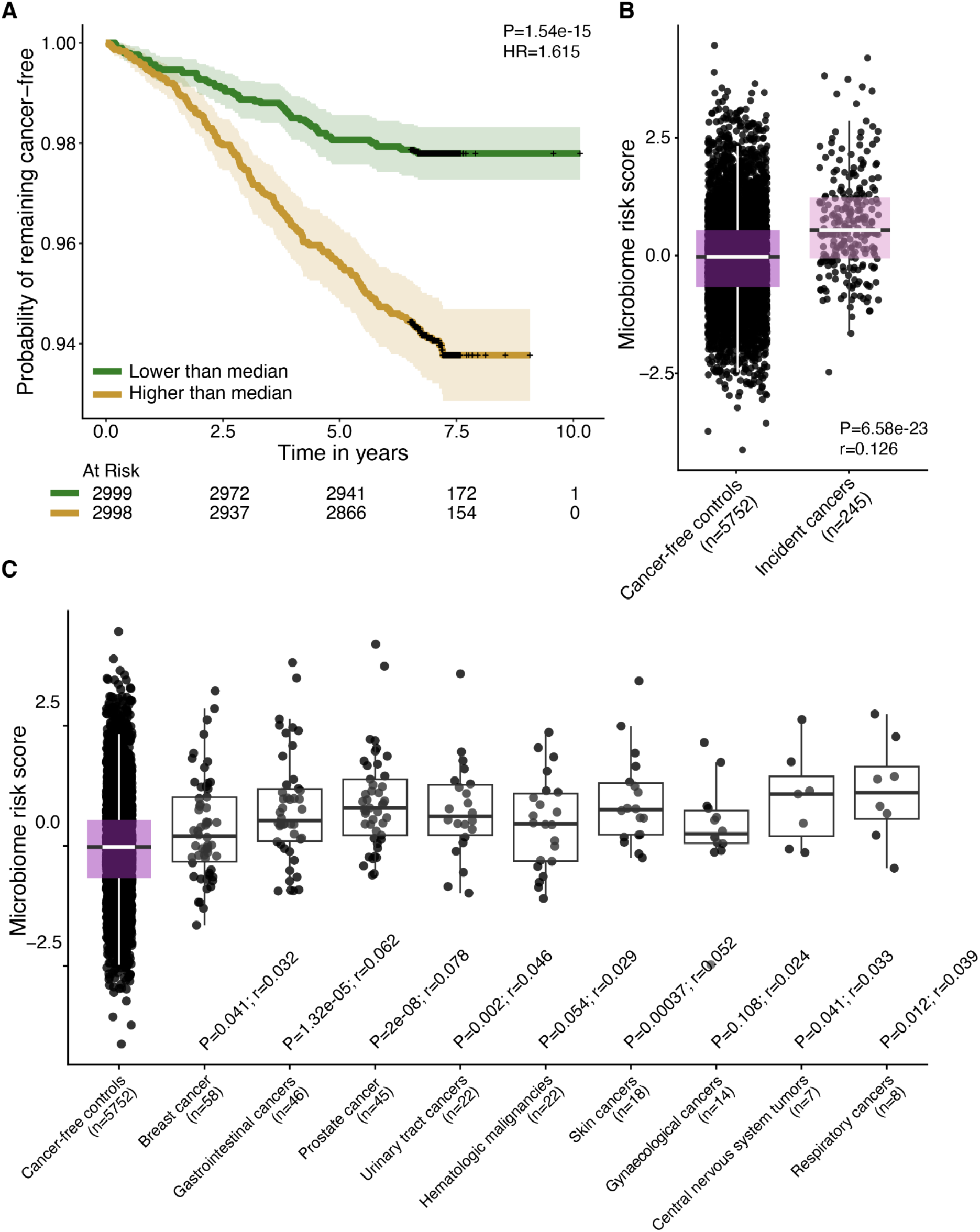
Microbiome risk score is associated with future incident cancer. **(A)** Kaplan-Meier curves showing cancer-free survival stratified by the median microbiome risk score. Individuals with a microbiome risk score above the median (yellow line) exhibited lower cancer-free survival during follow-up than individuals with a score below the median (green line), corresponding to a higher incidence of cancer. Shaded areas represent 95% confidence intervals. The reported P value was derived from a multivariable Cox proportional hazards model that included the microbiome risk score as a continuous variable and was adjusted for age, sex, BMI, smoking, baseline disease status, the Lifelines Diet Score, and Bristol Stool Scale. The hazard ratio (HR) corresponds to the effect of 1-SD increase in the microbiome risk score. Tick marks indicate censored observations, and the numbers at risk are shown below the plot. **(B)** Distribution of the microbiome risk score in individuals who remained cancer-free and those who developed incident cancer during follow-up. P value was calculated using a one-sided Wilcoxon rank-sum test. **(C)** Distribution of the microbiome risk score according to the type of cancer diagnosed during follow-up. Cancer types are ordered by sample sizes which are indicated below each category. P values represent one-sided Wilcoxon rank-sum tests comparing individuals with each cancer type with cancer-free individuals and were adjusted for multiple comparisons using the Benjamini-Hochberg false discovery rate procedure. Skin cancers are excluding basal cell carcinoma and squamous cell carcinoma. For (B-C) the corresponding Wilcoxon effect sizes (r) are shown; positive values indicate higher scores among individuals with cancer. We excluded bone/soft tissue cancers (n=3) and endocrine cancers (n=2) from panel C due to the low number of cases. However, these cases are included in panel A and B.

To determine whether the microbiome cancer-risk score reflected cancer-specific biology or a broader disease-related signature, we examined its association with non-cancer diseases at baseline as well as with the subsequent development of chronic disease. The microbiome risk score was not associated with non-cancer diseases at baseline (P=0.75; **Fig. S3A**), nor with incident cardiovascular disease (P=0.094; **Fig. S3A**) or immune-mediated inflammatory disease (P=0.39; **Fig. S3B**) self-reported during follow-up. The score was also not associated with subsequent development of obesity, comparing individuals who progressed from lean or overweight to obese with those who remained lean during follow-up (P=0.64; **Fig. S3C**). Only 18 of 245 individuals who developed cancer also developed another non-cancer disease during follow-up, indicating limited overlap between these incident disease groups (**Fig. S4**). Together, these findings suggest that the cancer-associated microbiome signature is not simply a marker of prevalent disease or a general predisposition to disease.

To evaluate the external generalizability of our microbiome cancer-risk score, we applied it to independent metagenomic datasets comprising 1,879 patients with prevalent cancers and 5,341 cancer-free individuals from 36 published studies^23^, as well as to the independent population-based FINRISK cohort, which included 328 individuals with incident cancer and 5,636 individuals without cancer during follow-up. Follow-up in FINRISK was limited to 10 years after stool sampling to match Lifelines. Because information on premalignant or benign lesions was unavailable in FINRISK, we additionally performed a two-year landmark analysis to reduce the likelihood that the microbiome cancer-risk score reflected undiagnosed or preclinical disease present at stool sampling. Participants who developed cancer, died, or were lost to follow-up during the first two years were excluded, and survival follow-up began at the two-year landmark.

Across the external case-control datasets, patients with established cancer had higher microbiome cancer-risk scores than cancer-free individuals (P=3.22e-07; **Fig. 2A**). This pattern was observed across multiple cancer types present at the time of stool sampling, with the greatest separation for colorectal, kidney and lung cancers (**Fig. 2B**). Applying the microbiome cancer-risk score to FINRISK without re-training revealed visible separation of the Kaplan-Meier curves between individuals with high (above median) and low (below median) cancer-risk scores approximately 1-2 years after the two-year landmark, corresponding to 3-4 years after stool sampling (**Fig. 2C**). This separation was, however, not sustained outside this interval, and the overall difference across the eight years of post-landmark follow-up was not statistically significant (**Fig. 2C**).

**Figure 2.**
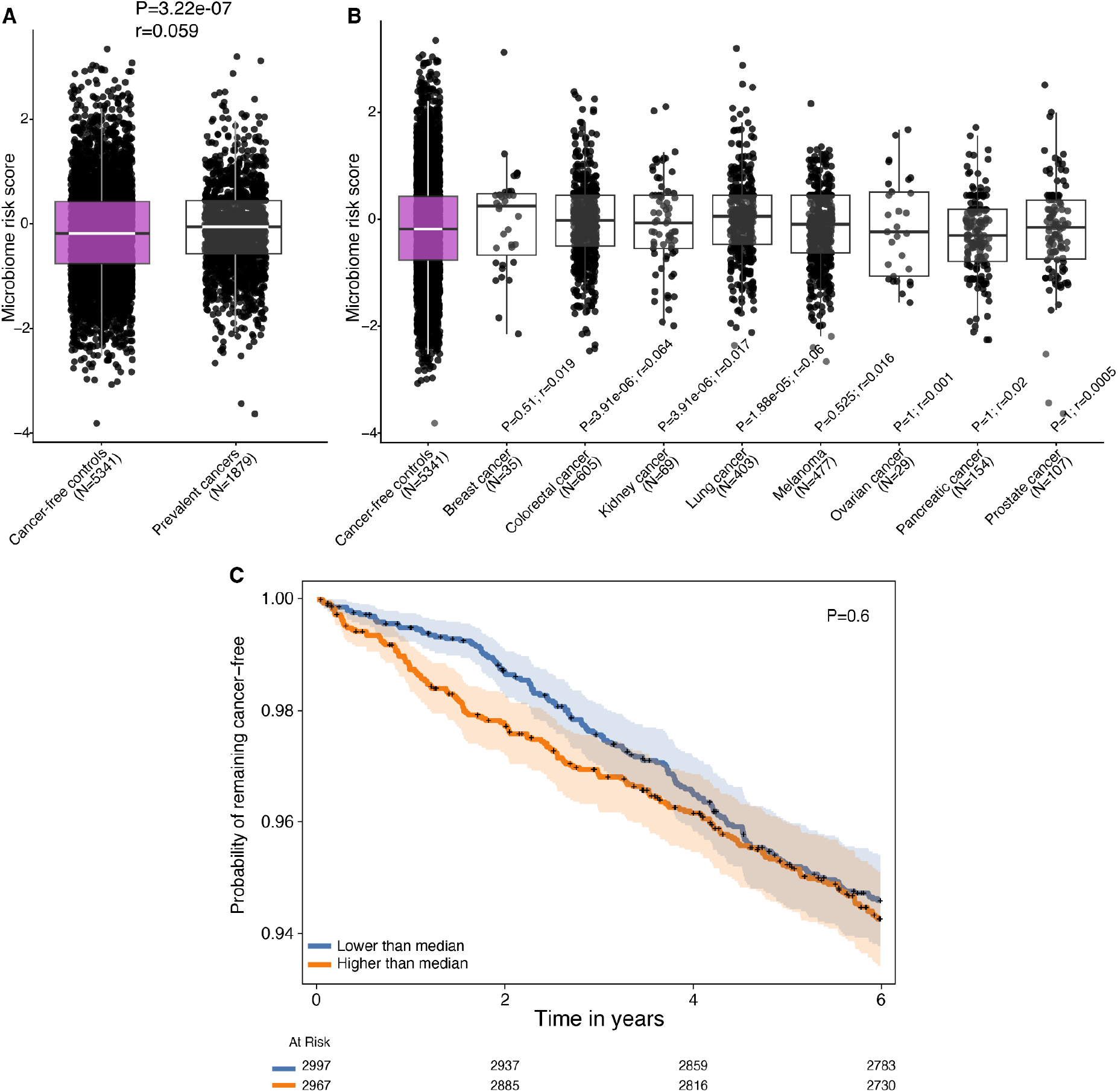
External validation of the microbiome risk score in independent prevalent and incident cancer cohorts. **(A)** Distribution of the microbiome risk score in participants with prevalent cancer and cancer-free controls across independent external metagenomic datasets. P values were calculated using one-sided Wilcoxon rank-sum tests. **(B)** Cancer types are ordered by sample sizes which are indicated below each category. P values represent one-sided Wilcoxon rank-sum tests comparing individuals with each cancer type with cancer-free individuals and were adjusted for multiple comparisons using the Benjamini-Hochberg false discovery rate procedure. **(C)** Kaplan-Meier curves showing the probability of remaining cancer-free in the independent FINRISK cohort, stratified by the median microbiome risk score. Participants were followed from two years after baseline microbiome sampling to minimize the influence of occult cancer at enrolment. Shaded areas represent 95% confidence intervals. Tick marks indicate censored observations, and the numbers at risk are shown below the plot.

Interpretation of the FINRISK findings should consider important differences between the two cohorts, including geographic location and population ancestry, dietary and lifestyle characteristics, cancer ascertainment and case definitions, as well as technical differences in stool processing, DNA extraction protocols, and sequencing depth. These sources of biological and technical heterogeneity may influence the observed microbiome composition and reduce the transferability of microbiome-derived risk models across cohorts.

The microbiome cancer-risk score comprised SGBs with both positive and negative associations with future cancer risk, indicating that the predictive signal was distributed across multiple microbial taxa rather than dominated by a single organism. Several taxa represented in the cancer-risk score, or their broader taxonomic groups, have previously been implicated in cancer (**Table S2**). For example, *Candidatus Parachristensenella avicola* (SGB58519) was enriched in melanoma patients with poor response to immune checkpoint inhibitors^22^, while *Desulfovibrio piger* (SGB15467), a member of the hydrogen sulfide-producing Desulfovibrionaceae, has been associated with multiple prevalent cancers^25^. *Sutterella wadsworthensis* (SGB9283) and a *Lachnospiraceae* species (SGB4782), both negatively associated with cancer incidence, were associated with treatment response or cancer-free status in the Gut OncoMicrobiome Signatures (GOMS) meta-analysis^23^ (**Table S2)**.

Although some cancer-score-associated SGBs were phylogenetically close to one another, those associated with higher and lower risk did not form distinct clades but were instead interspersed across the phylogenetic tree **(Fig. S5)**. To place unclassified SGBs in a phylogenetic context, we identified their nearest named relatives **(Fig. S5)**. Two additional unclassified SGBs positively associated with cancer incidence were most closely related to *Dysosmobacter welbionis* and *Flavonifractor plautii*, respectively, both of which were also reported in the GOMS meta-analysis^23^. Other SGBs in the cancer-risk score have not previously been linked to cancer and may therefore represent novel candidate biomarkers. The score did not include canonical pathobionts commonly associated with gut dysbiosis, such as members of the *Enterocloster* genus^23^. This may depend on multiple factors, including variation in gut microbial signatures across stages of cancer development^14^ and the heterogeneous composition of cancer types in our cohort. Differences in tumour immunogenicity could also influence the extent to which the gut microbiome contributes to disease development^26^.

This study has several limitations. First, gut microbiome sampling was performed at a single time point, precluding assessment of the temporal stability of the microbiome risk score. Demonstrating long-term stability in individuals who remain cancer-free would strengthen the interpretation that the score reflects a persistent host-microbiome phenotype associated with future cancer risk rather than an early response to occult tumour development. Second, cancer cases were identified through linkage with Palga and required histological confirmation, providing high diagnostic accuracy. However, controls were defined by the absence of histologically confirmed cancer before sampling or during follow-up. Consequently, occult or undiagnosed cancer at the time of stool sampling cannot be excluded. Third, although the microbiome risk score was developed using all incident cancer cases combined, the number of cases for individual cancer types was insufficient to derive or validate cancer type-specific signatures. Nevertheless, the distribution of cancer types within the cohort reflected population-level cancer incidence in the Netherlands, supporting the representativeness of the study population^27^.

In conclusion, we identified a gut microbiome-derived risk score associated with subsequent cancer diagnosis, in some cases several years after microbiome sampling, and evaluated its generalizability in independent prevalent and incident cancer cohorts. The overlap with microbial signatures observed in established cancer suggests that cancer-associated microbial alterations may already be detectable before clinical diagnosis and could serve as early biomarkers of future cancer risk. Larger longitudinal cohorts with repeated microbiome sampling and mechanistic studies are needed to determine whether these alterations contribute causally to carcinogenesis or primarily reflect underlying cancer risk or early, undiagnosed disease.

## Methods

### Study cohort

We analyzed 5,997 adult participants from the Lifelines Biobank with available stool metagenomic profiles from the Dutch Microbiome Project (DMP), linked to histologically confirmed cancer diagnoses in the Dutch Nationwide Pathology Databank (Palga). Cancer diagnoses occurring before stool sampling, premalignant or benign lesions, uncertain cancer diagnoses and self-reported previous cancer without a corresponding Palga record were excluded (**Fig. S1)**. Participants were followed from stool sampling (between January 2013 to September 2016) until cancer diagnosis or the end of available Palga follow-up (16 March 2023), resulting in individual follow-up periods of up to 10 years. During follow-up, 245 participants developed incident cancer and 5,752 remained cancer-free. Clinical and demographic features, including age, sex, BMI, current medication use, diet FFQ and Bristol Stool Scale were assessed at time of stool sampling as a part of the Lifelines study^21^. A subset of participants underwent a second assessment between 2019-2023, comprising questionnaires and anthropometric measurements. We used this follow-up assessment to identify incident cardiovascular disease (CVD), immune-mediated inflammatory disease (IMID) and obesity.

### Patient inclusion

Participants in the Dutch Microbiome Project were linked to Palga through March 16, 2023 (study end date), enabling identification of cancer diagnoses and their histopathological characteristics. Palga pathology reports contain extensive histopathological information, including tumor subtype, size, anatomical location, and cancer stage. Cancer cases were grouped according to the classification system of the Dutch Integrated Cancer Centre (IKNL) and by time of diagnosis.

Pre-malignant and benign lesions, including cervical dysplasia, vulvar leukoplakia, ductal carcinoma in situ, Barrett’s esophagus, basal cell carcinoma, and cutaneous squamous cell carcinoma, as well as uncertain cancer diagnoses, were excluded. Participants with a cancer diagnosis recorded in Palga before microbiome sampling or a self-reported history of cancer in Lifelines were excluded. Incident cancer cases were defined as the first histologically confirmed cancer diagnosis recorded in Palga after microbiome sampling, resulting in 245 participants with incident cancer available for analysis (Fig. S1).

The Lifelines study was approved by the medical ethical committee of the University Medical Center Groningen (METc number: 2017/152). Additional written consent was signed by all DMP participants.

### Stool sample collection, DNA extraction and sequencing

Stool samples were collected and processed as described by Gacesa et al^10^. Stool samples were collected between January 2013 and 2016. Participants collected stool samples at home and froze them within 15 minutes of defecation. Frozen samples were collected by Lifelines, transported on dry ice, and stored at −80°C at the University Medical Center Groningen (UMCG). DNA was isolated using the QIAamp Fast DNA Stool Mini Kit (Qiagen) on the QIAcube automated sample preparation system (Qiagen). Samples with DNA yields below 200 ng (measured using a Qubit 4 Fluorometer) were prepared using the NEBNext Ultra DNA Library Prep Kit for Illumina, whereas all other samples were prepared using the NEBNext Ultra II DNA Library Prep Kit for Illumina. Metagenomic sequencing was performed by Novogene on the Illumina HiSeq 2000 platform, generating approximately 8 Gb of 150-bp paired-end reads per sample (mean 7.9 Gb, SD 1.2 Gb).

### Metagenomic data processing

Illumina adapters and low-quality bases were trimmed with Trimmomatic as implemented in KneadData v0.12.4, using a leading and trailing base-quality threshold of Phred 20 and a 4-base sliding window requiring mean Phred ≥ 20. Reads shorter than 50 bp after trimming were discarded. Reads mapping to the human reference genome (GRCh37/hg19) were removed with Bowtie2 v2.5.1 in --very-sensitive end-to-end mode, with any aligning read discarded. Read quality was assessed with FastQC v0.12.1 before and after processing. Taxonomic profiles were generated using MetaPhlAn4 with the mpa_vOct22_CHOCOPhlAnSGB_202212 database using default parameters.

### Statistical analyses

#### Development of the gut microbiome risk score

Model development used repeated stratified random-split validation, comprising 100 independent splits of the study population into training (70%) and test (30%) sets while preserving the proportions of participants who were subsequently diagnosed with cancer and those who remained cancer-free. Within each resampling iteration, microbial feature selection and model fitting were performed using the training set, and model performance was evaluated in the corresponding held-out test set. The modelling procedure was evaluated using both SGB presence-absence profiles and centered log-ratio (CLR)-transformed relative abundances.

Within each training set, SGBs were ranked according to the log ratio of their prevalence among participants who were subsequently diagnosed with cancer to their prevalence among participants who remained cancer-free. Prevalence was calculated after converting the feature table to presence-absence profiles. Candidate SGBs were required to have a prevalence of at least 10% in either group and to be present in at least five cancer-free participants, the latter criterion serving to stabilize the denominator of the prevalence ratio. A pseudocount of 0.01 was added to both prevalence estimates before calculating the log prevalence ratio. Up to 100 SGBs with the largest positive and 100 with the largest negative log prevalence ratios were retained, yielding a maximum of 200 candidate predictors per training set.

For the presence-absence representation, an elastic-net penalized Cox proportional hazards model was fitted within each training set using the SGBs selected in that training set. For the CLR representation, a common feature basis was defined as the union of SGBs retained by prevalence-ratio screening across the 100 resampling iterations. This common basis ensured that CLR values were calculated relative to the geometric mean of the same set of SGBs in every iteration. CLR transformations were performed separately for the corresponding training and test feature tables using this common basis.

Elastic-net Cox models were fitted using α=0.5. The regularization parameter λ was selected by fivefold internal cross-validation within each training set, using Harrell’s concordance index as the optimization criterion (glmnet::cv.glmnet, type.measure=“C”); the value minimizing the cross-validation error (lambda.min) was retained. Raw microbiome risk scores were calculated as the model linear predictors. Within each resampling iteration, scores in both the training and test sets were standardized using the mean and standard deviation of the raw linear predictors in the training set. Consequently, hazard ratios represent the relative hazard associated with a one-standard-deviation increase in the microbiome risk score, with the standard deviation defined in the corresponding training set.

Model performance was evaluated in the held-out test sets using Harrell’s C-index and hazard ratios from Cox proportional hazards models. Risk-score associations were evaluated both without adjustment and after adjustment for sex, age, BMI, smoking history, Bristol Stool Scale, and Lifelines Diet Score. Missing continuous covariates in the training and test sets were imputed using medians estimated from the corresponding training set, and age and BMI were centered using training-set means. The presence-absence and CLR representations were further compared using models containing both risk scores and analyses of the predictive information unique to each representation. Presence-absence profiles showed better overall performance and retained more independent predictive information than CLR-transformed relative abundances and were therefore selected for construction of the final microbiome risk score. This is in line with other recent reports on microbiome-based classification using presence-absence of features^28^.

Stable microbial features were defined as SGBs with non-zero coefficients in a consistent direction in at least 90% of the resampling iterations. The final microbiome risk score was constructed by averaging the elastic-net coefficients of these stable SGBs across the 100 iterations and applying the resulting coefficients to the presence-absence profiles of the complete study population.

#### Association of the final microbiome risk score with incident cancer

After constructing the final microbiome cancer-risk score, we evaluated its association with subsequent cancer diagnosis in the complete study population using Cox proportional hazards regression. The score was standardized so that hazard ratios represented the relative hazard per 1-SD increase. Missing values for continuous covariates were imputed using the corresponding median calculated across the complete study population. Models were adjusted for age, sex, BMI, smoking history, prevalent disease status, Bristol Stool Scale, and Lifelines Diet Score. Hazard ratios and corresponding 95% confidence intervals were obtained by exponentiating the Cox regression coefficients and their Wald confidence intervals.

In a sensitivity analysis, the multivariable model was additionally adjusted for alcohol consumption. Daily alcohol intake was estimated by multiplying the energy-adjusted proportion of total energy derived from alcohol by total energy intake and dividing by 7 kcal/g. Alcohol consumption was categorized as high at >20 g/day for men and >10 g/day for women and as low otherwise. Participants with missing alcohol data were excluded from this analysis. The proportional-hazards assumption was assessed using scaled Schoenfeld residuals.

This was performed in R version 4.4.2 (2024-10-31) using tidyverse (v 2.0.0) and dplyr (v 1.2.0). The Elastic-net Cox models were fitted using the glmnet 4.1.10 and survival (v 3.8.3). CLR transformation was performed using the compositions::clr (v 2.0.9) and zCompositions::cmultRepl (v 1.5.0.5) was used for zero imputation. Rstatix::wilcox_test and wilcox_effectsize (v 0.7.3) were used for testing differences in group medians and calculating the effect size.

#### Construction of the phylogenetic tree

The 28 SGBs contributing to the microbiome cancer-risk score were placed in the MetaPhlAn 4 ChocoPhlAn reference phylogeny (mpa_vJan25_CHOCOPhlAnSGB_202503). We did not infer a new phylogeny. For SGBs without a species-level name, we identified the nearest named relatives by moving outwards from each tip through successive ancestral nodes, until the surrounding clade contained at least eight named species within the same phylum. Where no-named species were found within 20 nodes, we searched for the nearest named genus, up to 25 nodes. One SGB (SGB8599) has no-named relative at either rank and is therefore shown without a phylogenetic context. We then pruned the phylogenetic tree to these SGBs and their named relatives (100 tips) using ape::keep.tip, retaining the original branch lengths, and plotted it with ggtree. This analysis was performed in R 4.6.0 with ape (v 5.8-1), phangorn (v 2.12.1), ggtree 4.2.0, and ggplot2 (v 4.0.3).

### External validation

#### Established cancer vs cancer-free controls

We used publicly available stool metagenomic taxonomic profiles and accompanying metadata compiled for the Gut OncoMicrobiome Signatures (GOMS) pan-cancer metaanalysis^22^ (https://github.com/andrewmaltezthomas/NRCO_GOMS/tree/master/data/Cancer_vs_Controls_meta_analysis.rds). The compilation comprises 1,879 adults with eight different cancer types from 30 cohorts across 23 studies and 5,341 individuals without cancer from 17 cohorts across 14 studies. All samples had been taxonomically profiled at the species-level genome bin level using MetaPhlAn 4.

#### Independent incident cohort - FINRISK

Stool samples were obtained as part of FINRISK 2002, a national population survey based on a stratified random sample of Finnish adults aged 25-74 years from six geographical regions. Participants who agreed to take part were given a collection kit together with written instructions at the baseline visit and produced the sample at home. The samples were sent by overnight post in Finnish winter conditions to the Finnish Institute for Health and Welfare, held there at −20 °C, and shipped on dry ice to the University of California San Diego in 2017 for sequencing^29^.

Shotgun metagenomes were obtained for 7,231 participants using an Illumina HiSeq 4000 platform. Quality and adapter trimming was carried out with Atropos, after which reads of human origin were discarded by aligning to the GRCh38 assembly using Bowtie2. Taxonomic profiles were then generated from the remaining reads with MetaPhlAn v4.1.1 and its default reference database (mpa_vJun23_CHOCOPhlAnSGB_202403).

#### Application of the Lifelines microbiome cancer-risk score in independent cohorts

To evaluate whether the microbiome cancer-risk score generalized beyond Lifelines, we applied the final score without refitting to independent datasets comprising patients with established cancer and cancer-free controls, as well as to the prospective FINRISK cohort. For each external sample, a binary presence-absence profile was generated for the SGBs included in the score, with SGBs not detected in the external dataset assigned a value of zero. The linear predictor was calculated as the weighted sum of the SGB presence-absence values and their corresponding Lifelines coefficients. To place scores on the same scale as in the discovery cohort, external linear predictors were standardized using the mean and standard deviation of the linear predictor estimated in Lifelines.

In the established-cancer datasets, risk scores were compared between patients with cancer and cancer-free controls and across cancer types using Wilcoxon rank-sum tests. In FINRISK, associations between the standardized risk score and subsequent cancer diagnosis were evaluated using Kaplan-Meier curves. Analyses were performed using R version 4.4.2.

## Data Availability

All metagenomes analysed in this study will be made available upon publication. Metadata can be requested directly through Lifelines.

