## Supplementary information for "A gut microbiome-derived risk score is associated with future cancer development"

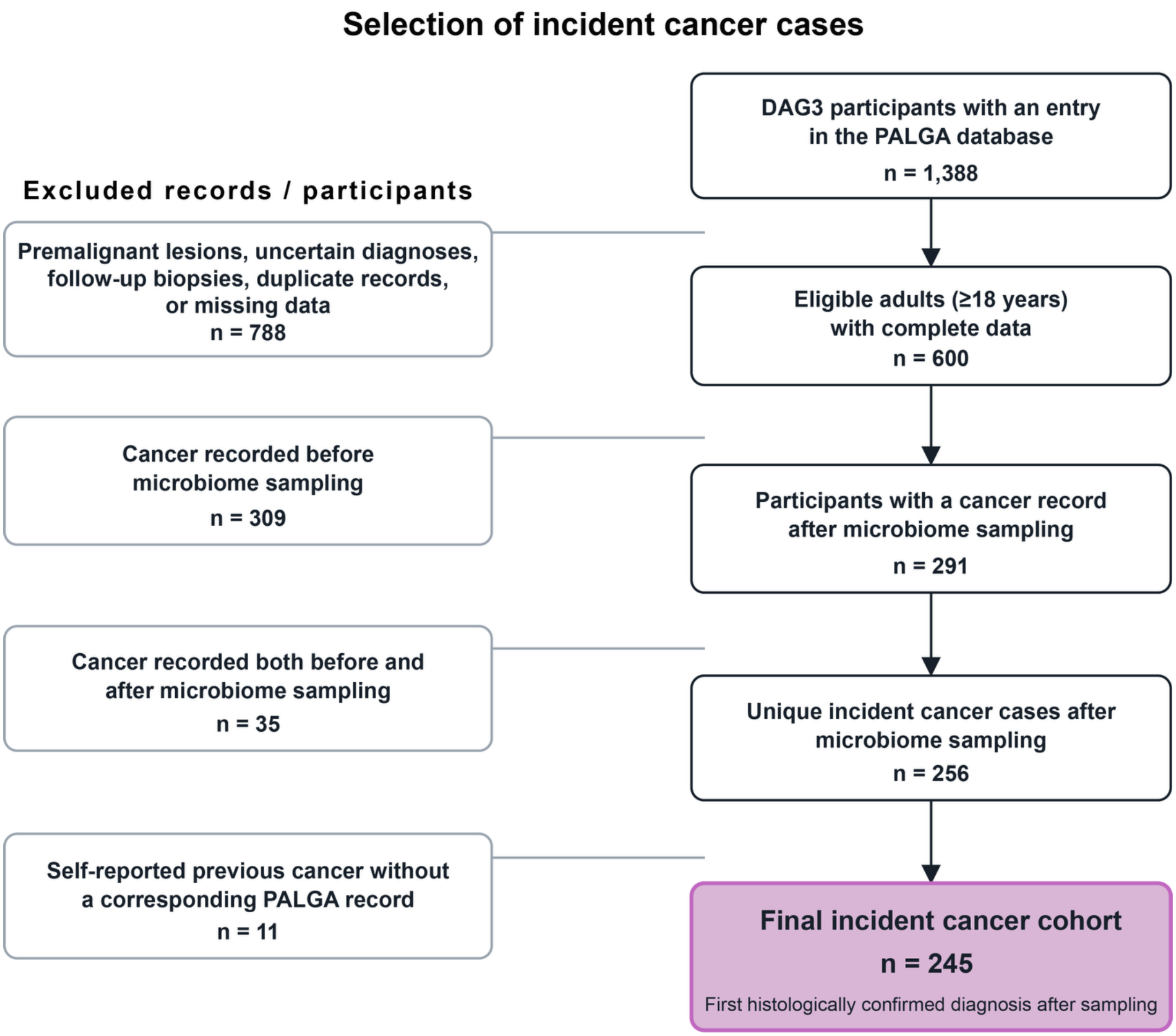


**Figure S1. Flow diagram of participant selection based on records from the Dutch Nationwide Pathology Databank (Palga).** Excluded Palga records comprised premalignant or benign lesions, uncertain diagnoses, follow-up biopsies, duplicate records, and records with missing metadata. Excluded conditions included cervical dysplasia, vulvar leukoplakia, ductal carcinoma in situ, Barrett's esophagus, basal cell carcinoma, and cutaneous squamous cell carcinoma. Participants were also excluded if cancer had been recorded before microbiome sampling or if they had self-reported a previous cancer diagnosis in Lifelines without a corresponding Palgarecord. Incident cancer was defined as the first histologically confirmed cancer diagnosis recorded in Palga after microbiome sampling.

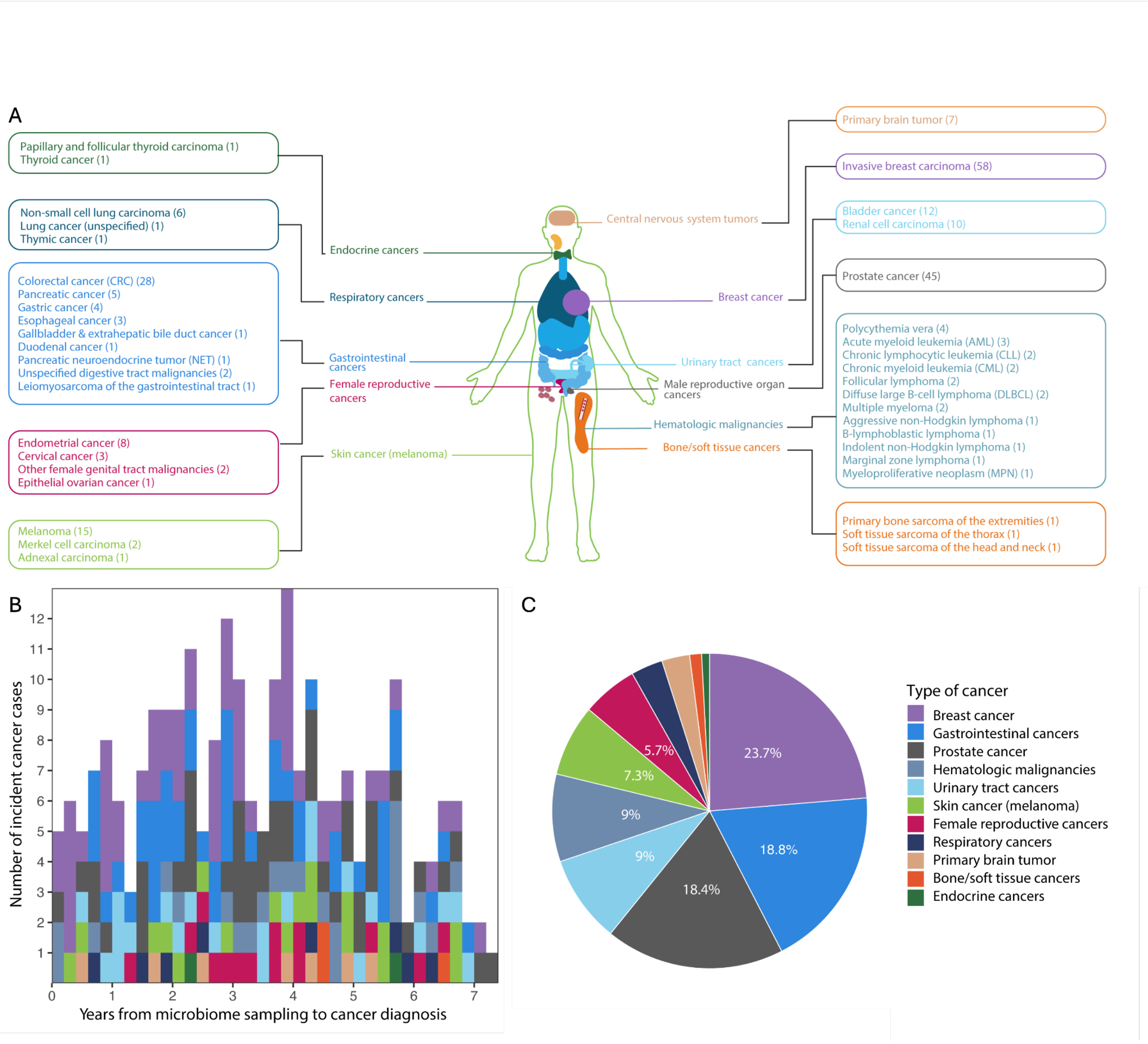


**Figure S2. Incident cancer diagnoses during follow-up.** (A) Incident cancer diagnoses (n=245 total) were grouped into 11 categories by organ system based on the tumour grouping from The Netherlands Cancer Registry (NCR)[1]. This grouping is based on ICD-O-3 topography and morphology codes. Each box lists the specific cancer subtypes diagnosed within that system, with the number of cases (n) in parentheses. (B) Number of incident cancer diagnoses colored by cancer type as a function of follow-up time (in years) from stool sampling to the study end date. (C) Proportional distribution of incident cancer diagnoses (n=245), per cancer type, coloured as in (B).

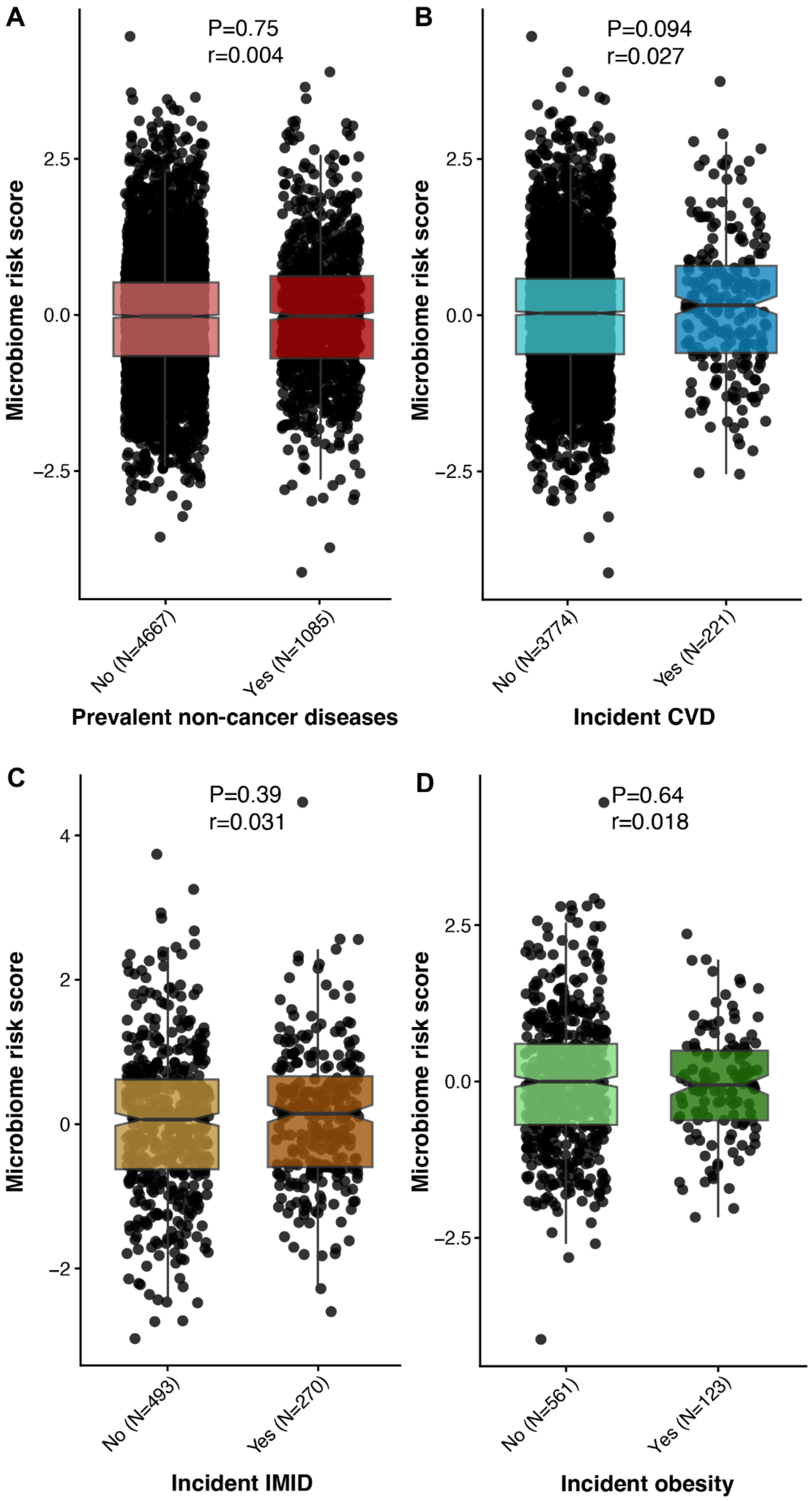

A
P=0.75
r=0.004
Microbiome risk score
2.5
0.0
−2.5
No (N=4667)
Yes (N=1085)
Prevalent non-cancer diseases
B
P=0.094
r=0.027
Microbiome risk score
2.5
0.0
−2.5
No (N=3774)
Yes (N=221)
Incident CVD
C
P=0.39
r=0.031
Microbiome risk score
4
2
0
−2
No (N=493)
Yes (N=270)
Incident IMID
D
P=0.64
r=0.018
Microbiome risk score
2.5
0.0
−2.5
No (N=561)
Yes (N=123)
Incident obesity

**Figure S3. Microbiome risk scores according to prevalent and incident disease status.** Microbiome risk scores are shown according to (A) prevalent non-cancer disease status at baseline and the development of (B) cardiovascular disease (CVD), (C) immune-mediated inflammatory disease (IMID), and (D) obesity after stool sampling. Incident obesity was defined as obesity at follow-up (BMI ≥30 kg/m²) among individuals who were lean (BMI <25 kg/m²) or overweight (BMI ≥25 and <30 kg/m²) at baseline. P values were calculated using two-sided Wilcoxon rank-sum tests.

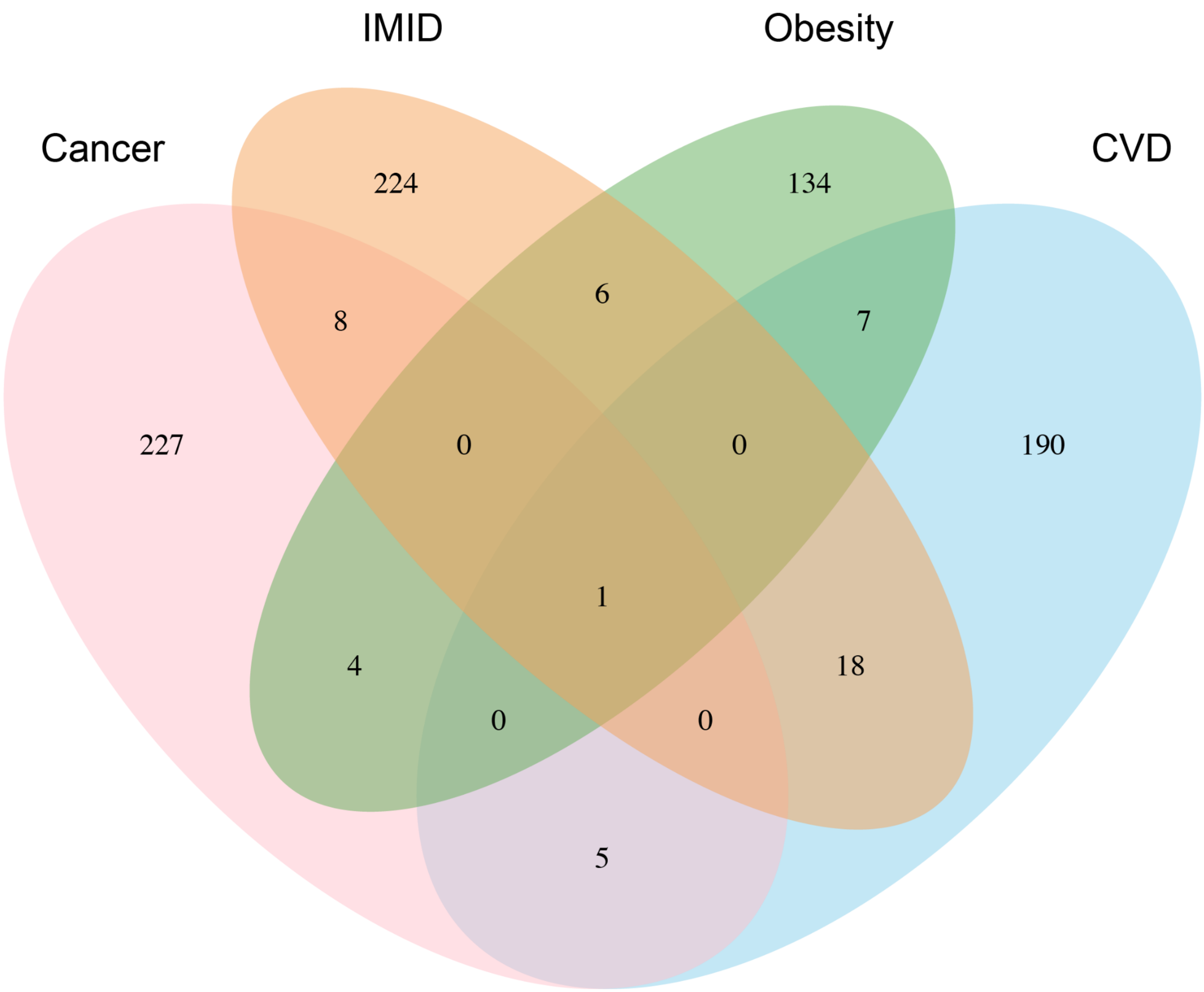


**Figure S4. Overlap among individuals with incident diseases.** Venn-diagram showing the number of individuals with one or multiple incident diseases, including incident cancer (pink), incident CVD (blue), incident IMID (orange) and incident obesity (green).

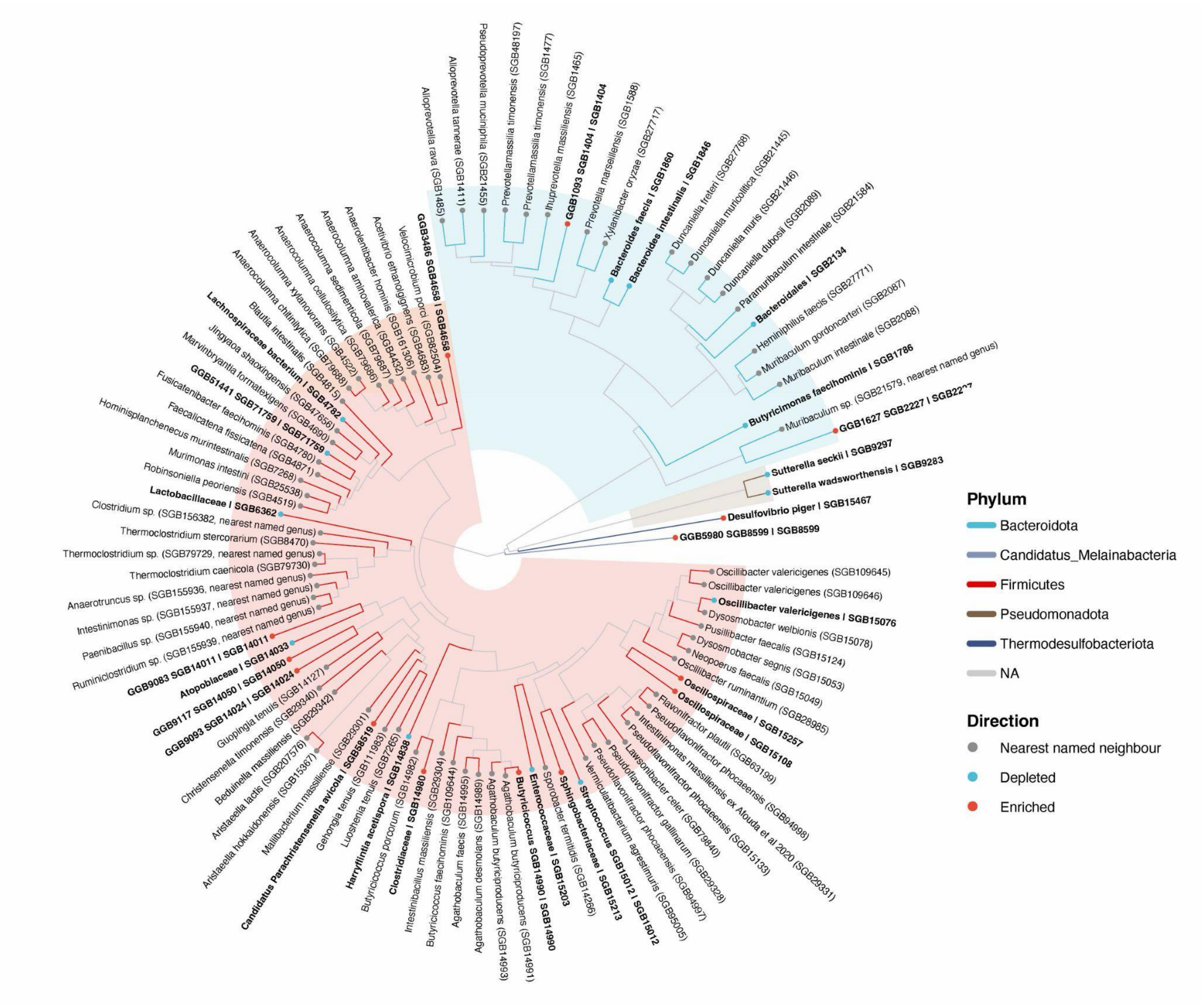


**Figure S5. Phylogenetic relationships of SGBs included in the microbiome risk score.** A phylogenetic tree (circular phylogram) including the 28 risk score SGBs and up to 8 closest relatives. SGBs that were positively and negatively associated with cancer risk are shown as red and blue nodes, respectively. Nodes in gray are their nearest named phylogenetic relatives. Branch colour and background shading indicate phylum. Labels marked "sp. (nearest named genus)" indicate that no named species was found within range of the focal risk score SGB. The tree was constructed from the ChocoPhlAn reference tree (mpa_vJan25_CHOCOPhlAnSGB_202503) **(see Methods)**.

**Table S1. Taxonomic classification and model coefficients of the 28 SGBs included in the microbiome risk score.** The table presents the order, family, species, and model coefficient for each SGB whose presence or absence contributed to the predicted risk of incident cancer. Red and blue cells indicate positive and negative associations with incident cancer risk, respectively.

| Order | Family | Species | SGB | Coefficient |
|---|---|---|---|---|
| *Eubacteriales* | *Clostridiaceae* | *Butyricicoccus* SGB14990 | SGB14990 | 0.536 |
| *Clostridia unclassified* | *Clostridia unclassified* | *Candidatus Parachristensenella avicola* | SGB58519 | 0.483 |
| *Eubacteriales* | *Clostridiaceae* | GGB9568 SGB14980 | SGB14980 | 0.423 |
| *Desulfovibronales* | *Desulfovibrionaceae* | *Desulfovibrio piger* | SGB15467 | 0.461 |
| *OFGB529* | *FGB529* | GGB1093 SGB1404 | SGB1404 | 0.396 |
| *OFGB658* | *FGB658* | GGB1627 SGB2227 | SGB2227 | 0.395 |
| *OFGB1422* | *FGB1422* | GGB3486 SGB4658 | SGB4658 | 0.352 |
| *OFGB2106* | *FGB2106* | GGB5980 SGB8599 | SGB8599 | 0.299 |
| *OFGB28372* | *FGB28372* | GGB9083 SGB14011 | SGB14011 | 0.328 |
| *OFGB2840* | *FGB2840* | GGB9093 SGB14024 | SGB14024 | 0.298 |
| *OFGB76428* | *FGB76428* | GGB9117 SGB14050 | SGB14050 | 0.269 |
| *Eubacteriales* | *Oscillospiraceae* | *Oscillospiraceae* unclassified | SGB15257 | 0.250 |
| *Eubacteriales* | *Oscillospiraceae* | GGB9636 SGB15108 | SGB15108 | 0.251 |
| *Sphingobacteriales* | *Sphingobacteriaceae* | GGB9697 SGB15213 | SGB15213 | 0.220 |
| *Lactobacillales* | *Streptococcaceae* | *Streptococcus* SGB15012 | SGB15012 | -0.182 |
| *Coriobacteriales* | *Atopobiaceae* | GGB9101 SGB14033 | SGB14033 | -0.193 |
| *Bacteroidales* | *FGB76185* | GGB1550 SGB2134 | SGB2134 | -0.244 |
| *Bacteroidales* | *Bacteroidaceae* | *Bacteroides faecis* | SGB1860 | -0.240 |
| *Bacteroidales* | *Bacteroidaceae* | *Bacteroides intestinalis* | SGB1846 | -0.239 |
| *Bacteroidales* | *Odoribacteraceae* | *Butyricimonas faecihominis* | SGB1786 | -0.244 |

| | | | | |
|---|---|---|---|---|
| *Lactobacillales* | *Enterococcaceae* | GGB33512 SGB15203 | SGB15203 | -0.279 |
| *OFGB9718* | *FGB9718* | GGB51441 SGB71759 | SGB71759 | -0.304 |
| *Eubacteriales* | *Oscillospiraceae* | *Harryflintia acetispora* | SGB14838 | -0.368 |
| *Eubacteriales* | *Lachnospiraceae* | *Lachnospiraceae bacterium unclassified* | SGB4782 | -0.344 |
| *Lactobacillales* | *Lactobacillaceae* | GGB4599 SGB6362 | SGB6362 | -0.356 |
| *Eubacteriales* | *Oscillospiraceae* | *Oscillibacter valericigenes* | SGB15076 | -0.397 |
| *Burkholderiales* | *Sutterellaceae* | *Sutterella seckii* | SGB9297 | -0.431 |
| *Burkholderiales* | *Sutterellaceae* | *Sutterella wadsworthensis* | SGB9283 | -0.475 |

**Table S2. Previously reported associations of SGBs included in the microbiome risk score**. For each SGB, the table summarizes previously reported associations involving the species or a higher taxonomic level with prevalent cancer (yellow), non-response (orange) or response (blue) to immune checkpoint inhibitor (ICI) therapy, or general health (green). The relevant study is cited, and the taxonomic level at which the association was reported is underlined.

| Order | Family | Genus (Species) | SGB | Prevalent cancer | ICI non-responders | ICI responders | General health |
|---|---|---|---|---|---|---|---|
| *Eubacteriales* | *Clostridiaceae* | *Butyricicoccus* SGB14990 | SGB14990 | | x Okazawa-Sakai et al. 2025 | | |
| *Clostridia unclassified* | *Clostridia unclassified* | *Candidatus Parachristensenella avicola* | SGB58519 | | x Björk et al. 2024, Alves Costa Silva et al. 2024 | | |
| *Eubacteriales* | *Clostridiaceae* | GGB9568 SGB14980 | SGB14980 | | | | x Humińska-Lisowska et al. 2024 |
| *Desulfovibronales* | *Desulfovibrionaceae* | *Desulfovibrio piger* | SGB15467 | x Yonekura et al. 2022 | | | |
| *OFGB529* | *FGB529* | GGB1093 SGB1404 | SGB1404 | | | | |
| *OFGB658* | *FGB658* | GGB1627 SGB2227 | SGB2227 | x Duttagupta et al. 2026 | | | |
| *OFGB1422* | *FGB1422* | GGB3486 SGB4658 | SGB4658 | | | | |
| *OFGB2106* | *FGB2106* | GGB5980 SGB8599 | SGB8599 | x Alves Costa Silva et al. 2024 | | | |

| | | | | | | | | | | | | |
|---|---|---|---|---|---|---|---|---|---|---|---|---|
| *OFGB28372* | *FGB28372* | GGB9083 SGB14011 | SGB14011 | | | | | | | | | |
| *OFGB2840* | *FGB2840* | GGB9093 SGB14024 | SGB14024 | | | | | | | | | |
| *OFGB76428* | *FGB76428* | GGB9117 SGB14050 | SGB14050 | | x | Duttagupta et al. 2026 | | | | | | |
| *Eubacteriales* | *Oscillospiraceae* | *Oscillospiraceae* unclassified | SGB15257 | | | | x | Nguyen et al. 2024 | | | | |
| *Eubacteriales* | *Oscillospiraceae* | GGB9636 SGB15108 | SGB15108 | | | | x | Nguyen et al. 2024 | | | | |
| *Sphingobacteriales* | *Sphingobacteriaceae* | GGB9697 SGB15213 | SGB15213 | | | | | | | | x | D'aiello et al. 2025 |
| *Lactobacillales* | *Streptococcaceae* | *Streptococcus* SGB15012 | SGB15012 | | x | Qu et al. 2023 | | | | | | |
| *Coriobacteriales* | *Atopobiaceae* | GGB9101 SGB14033 | SGB14033 | | x | Jimenez et al. 2025 | | | | | | |
| *Bacteroidales* | *FGB76185* | GGB1550 SGB2134 | SGB2134 | | | | | | x | Vetizou et al. 2015 | | |
| *Bacteroidales* | *Bacteroidaceae* | *Bacteroides faecis* | SGB1860 | | x | Mohebali et al. 2025 | | | | | | |
| *Bacteroidales* | *Bacteroidaceae* | *Bacteroides intestinalis* | SGB1846 | | | | x | Wang et al. 2009 | | | | |

| | | | | | | | | | | | |
|---|---|---|---|---|---|---|---|---|---|---|---|
| *Bacteroidales* | *Odoribacteraceae* | *Butyricimonas faecihominis* | SGB1786 | | | | | | | x | Yonekura et al. 2022 |
| *Lactobacillales* | *Enterococcaceae* | GGB33512 SGB15203 | SGB15203 | | | | | x | Matson et al. 2018, Guo et al. 2020 | | |
| *OFGB9718* | *FGB9718* | GGB51441 SGB71759 | SGB71759 | | | | | | | | |
| *Eubacteriales* | *Oscillospiraceae* | *Harryflintia acetispora* | SGB14838 | | | | | | | x | Asnicar et al. 2026 |
| *Eubacteriales* | *Lachnospiraceae* | *Lachnospiraceae bacterium unclassified* | SGB4782 | | | | | x | Kim et al. 2025, Thomas et al. 2023 | x | Gacesa et al. 2022, Pekel et al. 2026 |
| *Lactobacillales* | *Lactobacillaceae* | GGB4599 SGB6362 | SGB6362 | | | x | Colbert et al 2023 | | | | |
| *Eubacteriales* | *Oscillospiraceae* | *Oscillibacter valericigenes* | SGB15076 | | | | | | | x | Rosés et al. 2021 |
| *Burkholderiales* | *Sutterellaceae* | *Sutterella seckii* | SGB9297 | | | | | | | | |
| *Burkholderiales* | *Sutterellaceae* | *Sutterella wadsworthensis* | SGB9283 | | | | | | | x | Thomas et al. 2023 |

**Table S3.** Cohort characteristics at stool sample collection in FINRISK.

| Characteristic | Control<br>N = 5,636[1] | Case<br>N = 328[1] | p-value[2] | % Missing |
|---|---|---|---|---|
| **Age (years)** | -0.57 (12.76) | 9.77 (10.35) | <0.001 | 0.000 |
| **Sex** | | | 0.815 | 0.000 |
| 0 | 3,105 / 5,636 (55.1%) | 178 / 328 (54.3%) | | |
| 1 | 2,531 / 5,636 (44.9%) | 150 / 328 (45.7%) | | |
| **BMI ($kg/m^2$)** | -0.05 (4.67) | 0.79 (4.62) | <0.001 | 0.000 |
| **Smoking status** | | | 0.877 | 0.500 |
| 0 | 4,286 / 5,611 (76.4%) | 250 / 325 (76.9%) | | |
| 1 | 1,325 / 5,611 (23.6%) | 75 / 325 (23.1%) | | |
| **Chronic disease** | | | <0.001 | 0.000 |
| 0 | 4,100 / 5,636 (72.7%) | 195 / 328 (59.5%) | | |
| 1 | 1,536 / 5,636 (27.3%) | 133 / 328 (40.5%) | | |

[1] n / N (%); Mean (SD)

[2] Pearson's Chi-squared test; Wilcoxon rank sum test
